# Large Language Models for Supporting Clear Writing and Detecting Spin in Randomized Controlled Trials in Oncology

**DOI:** 10.1101/2025.05.05.25326991

**Authors:** Carole Koechli, Fabio Dennstädt, Christina Schröder, Daniel M. Aebersold, Robert Förster, Daniel R. Zwahlen, Paul Windisch

## Abstract

**Importance:** Accurate interpretation of randomized controlled trial (RCT) results is essential for guiding clinical practice in oncology. Reporting “spin” can misrepresent treatment efficacy, potentially leading to suboptimal clinical decisions. Standardized methods could help to detect misleading reporting.

**Objective:** To determine whether large language models (LLMs) can accurately classify oncology RCTs as positive or negative based on primary endpoint achievement when provided with different sections of the trial report, thereby assessing their utility in identifying potential spin in conclusions.

**Design:** Methodological evaluation using LLMs and human annotations of previously published clinical trials.

**Setting:** Random sample of RCTs from seven major medical journals published between 2005 and 2023.

**Participants:** 250 two-arm, single primary endpoint oncology RCT reports were randomly selected from the specified journals and publication years.

**Exposure(s):** Human annotators independently classified trials based on primary endpoint results before LLM evaluation. Three commercial LLMs (GPT-3.5 Turbo, GPT-4o, and o1) classified trials based on four different text inputs: 1) conclusion only, 2) methods and conclusion, 3) methods, results, and conclusion, or 4) title and full abstract.

**Main Outcome(s) and Measure(s):** Performance of LLMs in classifying trials as positive or negative, primarily measured using the F1 score.

**Results:** The analysis included 250 RCT reports; based on human annotation, 146 (58.4%) were positive and 104 (41.6%) were negative. o1 demonstrated the highest performance across all input conditions, achieving F1 scores of 0.932 (conclusion only), 0.960 (methods and conclusion), 0.980 (methods, results, and conclusion), and 0.970 (title and full abstract). Analysis of trials incorrectly classified as positive by the LLM when using only the conclusion revealed patterns such as absence of primary endpoint data, emphasis on secondary or subgroup findings, or unclear endpoint distinctions within the conclusion.

**Conclusions and Relevance:** LLMs can accurately classify oncology RCT outcomes. Discrepancies between classifications based on conclusions versus more complete text indicate potential spin. This approach could serve as a valuable supplementary tool for researchers, reviewers, and editors to enhance transparency and critical appraisal of oncology trial reporting, though further validation is required, especially for trials with more complex designs.

**Key Points:** *Question:* Can large language models (LLMs) accurately classify oncology randomized controlled trials (RCTs) as positive or negative based on primary endpoint achievement, and can this help identify potential “spin” in trial conclusions?

*Findings:* In this methodological evaluation of 250 oncology RCTs, the o1 LLM achieved high accuracy in classifying trials based on the title and full abstract, outperforming classifications based on the conclusion alone. Discrepancies between classifications using conclusions versus more complete text often indicated patterns that could be considered as spin.

*Meaning:* LLMs show promise as a supplementary tool to detect potential spin by identifying inconsistencies between conclusions and overall results.

## Introduction

Randomized controlled trials (RCTs) represent the gold standard for evaluating interventions in oncology.^1^ However, the reporting and interpretation of trial results can be subject to inconsistency and “spin” - the presentation of results in a way that may mislead readers about the true efficacy of interventions.^2^ This can, for example, be accomplished by emphasizing secondary endpoints or subgroup analyses when primary endpoints are not met. While most research that looked at the topic has found a substantial prevalence of spin, the exact number varies as it is not always straightforward to differentiate between what constitutes a balanced and comprehensive presentation of the results and what may be an attempt to mislead the reader.^3^

The growing capabilities of large language models (LLMs) could constitute a standardized way to determine the presence of spin. If an abstract is clearly written, a state-of-the-art LLM should be able to determine if its primary endpoint was met. As multiple studies have identified the conclusion as the most frequent source of spin^4,5^, we hypothesized that trials which are correctly classified as negative by an LLM when provided with the title and abstract, but incorrectly classified as positive when provided with only the conclusion, would be likely to contain some form of spin.

## Methods

Randomized controlled oncology trials from seven major medical journals (British Medical Journal, JAMA, JAMA Oncology, Journal of Clinical Oncology, Lancet, Lancet Oncology, New England Journal of Medicine) published between 2005 and 2023 were randomly sampled. To avoid edge cases for this feasibility study, the set of trials was further reduced to trial designs with exactly two arms and one primary endpoint.

Two authors (C.K. and P.W.) then manually classified 250 trials in a two-step process. After annotating the first twenty trials, all samples were discussed to recognize potential differences in the annotation criteria. The remaining trials were annotated separately and discrepancies were discussed after all trials had been annotated. A third author (D.R.Z.) was responsible to judge disagreements that persisted after discussion. However, since there were no cases of disagreement, this was not necessary. We used the abstract for annotation if it clearly stated the primary endpoint and its results; otherwise, we referred to the full publication or protocol. Three commercially available LLMs, namely Generative Pretrained Transformer (GPT) 3.5 Turbo, Generative Pretrained Transformer 4 Omni, and o1 (OpenAI, San Francisco, United States) were then tasked with classifying the trial as positive or negative. The three models were chosen to investigate whether the inherent capabilities of the model would impact their suitability for the classification task (e.g. simpler models requiring more explicit language to correctly identify trials). The respective model snapshots were gpt-3.5-turbo-0125, gpt-4o-2024-11-20, and o1-2024-12-17.The LLMs were called via the application programming interface (API) with the temperature parameter set to 1.

Each model was evaluated in four different rounds. In round one, the models were only provided with the conclusion of the abstract. In round two, the models were provided with the methods and conclusion of the abstract. In round three, the models were provided with the methods, results, and conclusion of the abstract. In round four, the models were provided with the title and the full abstract.

The following system prompt was used: *“You will be provided with the {section} of a randomized controlled oncology trial. Your task will be to classify if the trial was positive, i*.*e. if it met its primary endpoint, or negative, i*.*e. if it did not meet its primary endpoint. Your response should be either the word POSITIVE (in all caps) or NEGATIVE (in all caps)*.*”*

The “{section} was replaced with either “conclusion”, “methods and conclusion”, “methods, results, and conclusion”, or “title and abstract”. The user prompt was the respective title, abstract or section(s) of the abstract.

The results were evaluated against the ground truth by creating confusion matrices and computing several performance metrics (i.e. accuracy, precision, recall, and F1 score). 95% confidence intervals were estimated using normal approximation intervals. For the best performing model, we further analyzed and categorized the trials that were incorrectly predicted as positive when provided with the conclusion but were correctly predicted as negative when provided with the title and abstract.

All programming was performed in Python (version 3.13.2) using, among others, the pandas (version 2.2.3) and openai (version 1.67.0) packages.

## Results

Annotators disagreed on 2.8% of trials (n=7). All disagreements were caused by simple mistakes and could be easily resolved during the discussion. Ultimately, 58.4% (n=146) of trials were annotated as positive and 41.6% (n=104) as negative.

The performances of the models when provided with different sections of the abstract is depicted in Figure 1 and Table 1. o1 exhibited the best performance in each round with F1 scores of 0.932 (conclusion only), 0.96 (methods and conclusion), 0.98 (methods, results, and conclusion), and 0.97 (title and abstract). GPT-4o’s F1 scores across the four rounds were 0.89, 0.91, 0.94, and 0.94. GPT-3.5 Turbo exhibited F1 scores of 0.89, 0.92, 0.91, and 0.91.

**Table 1.** Classification performance. Accuracy, precision, recall and F1 score for GPT-3.5 Turbo, GPT-4o and o1 when predicting if a trial was positive based on different sections of the abstract. Numbers in parentheses indicate the 95% confidence intervals.

|  |  | Accuracy | Precision | Recall | F1-Score |
| --- | --- | --- | --- | --- | --- |
| Conclusion | GPT-3.5 Turbo | 0.87 (0.83 - 0.91) | 0.86 (0.82 - 0.90) | 0.93 (0.90 - 0.96) | 0.89 (0.86 - 0.93) |
|  | GPT-4o | 0.87 (0.83 - 0.91) | 0.91 (0.87 - 0.94) | 0.87 (0.83 - 0.91) | 0.89 (0.85 - 0.93) |
|  | o1 | 0.92 (0.89 - 0.95) | 0.92 (0.89 - 0.95) | 0.95 (0.92 - 0.97) | 0.93 (0.90 - 0.96) |
| Methods + conclusion | GPT-3.5 Turbo | 0.90 (0.86 - 0.94) | 0.90 (0.86 - 0.93) | 0.94 (0.91 - 0.97) | 0.92 (0.88 - 0.95) |
|  | GPT-4o | 0.90 (0.86 - 0.94) | 0.94 (0.91 - 0.97) | 0.88 (0.84 - 0.92) | 0.91 (0.88 - 0.95) |
|  | o1 | 0.95 (0.93 - 0.98) | 0.95 (0.93 - 0.98) | 0.97 (0.94 - 0.99) | 0.96 (0.93 - 0.98) |
| Methods + results + conclusion | GPT-3.5 Turbo | 0.90 (0.86 - 0.93) | 0.91 (0.88 - 0.95) | 0.91 (0.88 - 0.95) | 0.91 (0.88 - 0.95) |
|  | GPT-4o | 0.94 (0.91 - 0.97) | 0.99 (0.97 - 1.00) | 0.90 (0.87 - 0.94) | 0.94 (0.91 - 0.97) |
|  | o1 | 0.97 (0.95 - 0.99) | 0.98 (0.96 - 1.00) | 0.97 (0.95 - 0.99) | 0.98 (0.96 - 0.99) |
| Title + abstract | GPT-3.5 Turbo | 0.90 (0.86 - 0.93) | 0.90 (0.86 - 0.94) | 0.92 (0.89 - 0.96) | 0.91 (0.88 - 0.95) |
|  | GPT-4o | 0.93 (0.90 - 0.96) | 0.96 (0.94 - 0.99) | 0.91 (0.88 - 0.95) | 0.94 (0.91 - 0.97) |
|  | o1 | 0.97 (0.95 - 0.99) | 0.98 (0.96 - 1.00) | 0.97 (0.94 - 0.99) | 0.97 (0.95 - 0.99) |

**Figure 1.**
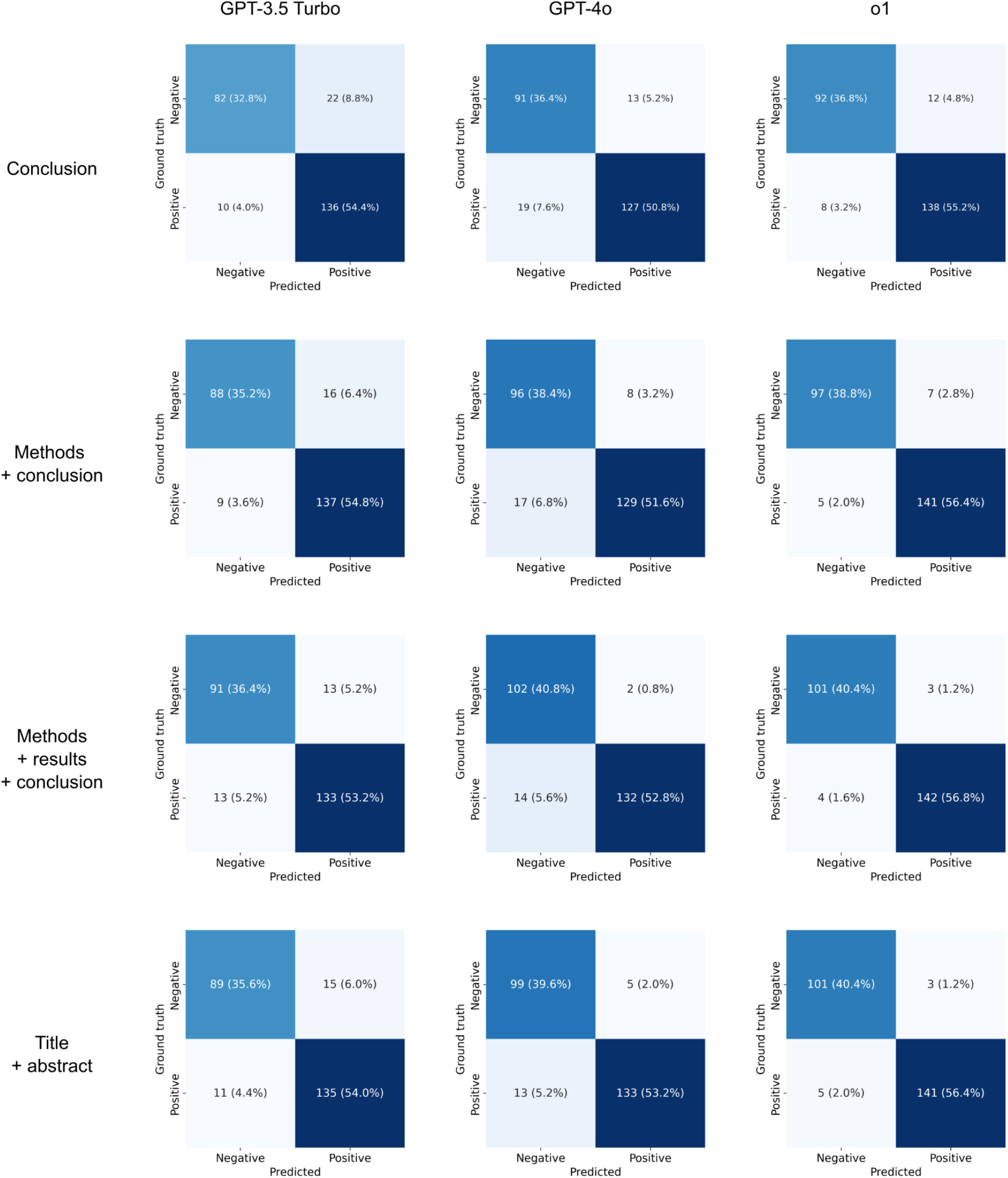
Confusion matrices. Classification performance of GPT-3.5 Turbo (left), GPT-4o (middle), and o1 (right) when predicting if a trial was positive or negative based on different sections of the abstract.

We further analyzed trials that were incorrectly predicted as positive by o1 when the model was only provided with the conclusion but predicted correctly when provided with the title and abstract. Of these ten trials, six did not mention the primary endpoint in the conclusion.^6–11^ One mentioned an improvement of the primary endpoint in a subgroup.^12^ One mentioned both improved secondary endpoints and the unimproved primary endpoint without specifying which was which.^13^ The remaining two trials mentioned that one arm was superior to the other one without specifying that it was the control arm which showed improved results.^14,15^ The list of trials is provided in Table 2.

**Table 2.** Trials that were incorrectly predicted as positive by o1 when the model was only provided with the conclusion but predicted correctly when provided with the title and abstract.

| Title | Conclusion reports on primary endpoint | Possible reason for incorrect prediction |
| --- | --- | --- |
| Total Body Irradiation or Chemotherapy Conditioning in Childhood ALL: A Multinational, Randomized, Noninferiority Phase III Study | Yes | Conclusion mentions that total body irradiation (TBI) plus etoposide showed improved overall survival. The model therefore likely thought that TBI plus etoposide was the intervention that was tested while it actually was the control. |
| Volasertib Versus Chemotherapy in Platinum-Resistant or -Refractory Ovarian Cancer: A Randomized Phase II Groupe des Investigateurs Nationaux pour l'Etude des Cancers de l'Ovaire Study | No | Primary endpoint not discussed in the conclusion. |
| High-Dose Therapy and Autologous Blood Stem-Cell Transplantation Compared With Conventional Treatment in Myeloma Patients Aged 55 to 65 Years: Long-Term Results of a Randomized Control Trial From the Group Myelome-Autogreffe | Yes | Conclusion mentions both improved secondary endpoints and the unimproved primary endpoint without specifying which is which |
| Results of a Randomized Trial of Chlorambucil Versus Fludarabine for Patients With Untreated Waldenström Macroglobulinemia, Marginal Zone Lymphoma, or Lymphoplasmacytic Lymphoma | No | Primary endpoint not mentioned in the conclusion. |
| Bortezomib-Dexamethasone, Rituximab, and Cyclophosphamide as First-Line Treatment for Waldenström's Macroglobulinemia: A Prospectively Randomized Trial of the European Consortium for Waldenström's Macroglobulinemia | No | Primary endpoint not mentioned in the conclusion. |
| Adjuvant tamoxifen and exemestane in early breast cancer (TEAM): a randomised phase 3 trial | No | Primary endpoint not mentioned in the conclusion. |
| Addition of Bevacizumab to Bolus Fluorouracil and Leucovorin in First-Line Metastatic Colorectal Cancer: Results of a Randomized Phase II Trial | No | Primary endpoint not mentioned in the conclusion. |
| Oral ibandronic acid versus intravenous zoledronic acid in treatment of bone metastases from breast cancer: a randomised, open label, non-inferiority phase 3 trial | Yes | Conclusion mentions superiority of zoledronic acid. The model therefore likely thought that zoledronic acid was the intervention, while it was the comparator in this non-inferiority study. |
| Bcl-2 Antisense (oblimersen sodium) Plus Dacarbazine in Patients With Advanced Melanoma: The Oblimersen Melanoma Study Group | Yes | Mentions improvement in the primary endpoint in a subgroup. |
| Efficacy and Safety of Trabectedin or Dacarbazine for Metastatic Liposarcoma or Leiomyosarcoma After Failure of Conventional Chemotherapy: Results of a Phase III Randomized Multicenter Clinical Trial | No | Primary endpoint not mentioned in the conclusion. |

## Discussion

In this study, we evaluated the ability of three commercial large language models to classify oncology randomized controlled trials as positive or negative based on different sections of trial abstracts. Our findings demonstrate that modern LLMs, particularly more advanced models, can achieve high classification accuracy even when provided with limited information. Our findings also support the hypothesis that trials which are correctly classified as negative by an LLM when provided with the title and abstract, but incorrectly classified as positive when provided with only the conclusion are likely to contain patterns that may be interpreted as spin. While there is no ground truth of what constitutes spin, not mentioning the results for the primary endpoint at all in the conclusion, mentioning an improvement of the primary endpoint that only occurred in a subgroup, or mixed reporting of primary and secondary endpoints without clear distinction would be at least considered questionable by many readers.^16^ Our findings also highlight that the LLM-based approach is not perfectly specific: Two of the ten studies’ conclusions clearly mentioned which arm had better outcomes. However, the LLM did not know which arm was the intervention and which arm was the control, so it assumed that the superior arm was the intervention arm. While this way of phrasing a conclusion may not be optimal for readability, it is certainly not an attempt at misleading the reader who will still know which treatment yielded better results.

Therefore, our approach is likely not suitable as a fully automated solution. However, it demonstrated its potential to inform editors, reviewers, and authors alike of potential spin or unclear writing. The question “Are the results for the primary endpoint clearly recognizable in the conclusion?” might serve as an alternative litmus test. Even though reviewers and journal editors are generally capable of recognizing questionable conclusions, we do believe that automated tools have value considering the ever-increasing list of items that have to be considered when conducting a careful review as they may, if implemented carefully, point towards parts of the manuscript that need increased attention.

A limitation of this study is that it is limited to RCTs with two arms and one endpoint. Multiple, primary or coprimary endpoints were not considered as this would have introduced additional complexity, e.g. by trying to identify if the alpha was appropriately split between endpoints.^17^ In addition, this study did not include designs other than standard hypothesis testing, such as Bayesian approaches.^18^

In conclusion, LLMs can effectively detect potential spin in oncology RCT reporting by identifying discrepancies between conclusions and full abstracts with high accuracy. This approach could serve as a supplementary tool for improving transparency in scientific reporting, though further development is needed to address complex trial designs beyond the two-arm, single-endpoint studies examined in this feasibility study.

## Data Availability

The data is available on github

https://github.com/windisch-paul/positive_negative

